# AI Generated Stromal Biomarkers for DCIS Reccurence Prediction

**DOI:** 10.64898/2026.02.13.26346278

**Authors:** Matthew McNeil, Vishwesh Ramanathan, Dina Bassiouny, Sharon Nofech-Mozes, Eileen Rakovitch, Anne L. Martel

## Abstract

**Background:** Although DCIS has a relatively low recurrence rate, many patients still receive adjuvant radiotherapy or endocrine therapy, raising concerns about overtreatment. Reliable biomarkers are therefore needed to predict an individual patient’s risk and guide treatment decisions. Recent studies suggest that the composition of the tumour-associated stroma (TAS) affects progression and outcome, highlighting TAS-derived biomarkers as promising candidates for further investigation.

**Methods:** We trained AI models for cell and tumour segmentation using whole slide digital pathology images acquired as part of a retrospective cohort study. We investigated the effects of cell density within both the tumour and the TAS to determine how they correlated with recurrence in the ipsilateral breast.

**Results:** We found that the concentration of DCIS lesions on the slide and the density of mitotic figures inside the TAS region were significantly associated with recurrence risk. Additionally, we found some predictive value in the lymphocyte and red blood cell densities in different tumour regions. Stromal composition was shown to associate with recurrence risk, and density-based biomarkers were identified and used to cluster patients into phenotypes with significantly different risk profiles.

**Conclusion:** Our findings highlight the prognostic relevance of stromal composition in DCIS, and we identify novel density-based biomarkers that can be used to identify patients who are more likely to experience a local recurrence after breast-conserving surgery alone. These results may aid in developing future risk-stratification tools for breast cancer patients, thereby reducing overtreatment and improving patient care.

## 1 Introduction

Due to the widespread use of mammography screening, ductal carcinoma in situ (DCIS) has become one of the most prevalently diagnosed types of cancer, representing 20-25% of all breast cancer cases [1]. Fortunately, patient prognosis is typically far better than for other cancer types, with a 10-year local recurrence rate of 30% for patients receiving breast-conserving surgery (BCS) alone[2][3][4]. It has been shown that this recurrence rate can be significantly reduced with the addition of radio- or endocrine-therapy to a patient’s treatment plan[5][6]. Despite this apparent benefit, recent work has shown that this may represent significant over-treatment for women with lower risk lesions[7][8]. Because of this, there is a need for biomarkers that can identify patients at a higher risk of recurrence. It is hoped that these could be used to better stratify patients into two groups: those who would benefit from more aggressive treatment, and those who could avoid its potential harmful effects without a significant risk of recurrence.

To identify these, some studies have achieved success using clinical variables, such as tumour size, margin status, and patient age[9]. Some efforts have been made to see if common histopathological indicators of disease prognosis in DCIS’ invasive counterpart, such as nuclear grade and necrosis, could have predictive value in DCIS, though with inconsistent success[9][10][5][11]. Commercial genomic assays such as the Oncotype DX DCIS Score have also been explored[12]; however, these tests are relatively expensive and require centralized laboratory processing, which can introduce delays into treatment decision-making. Given the limited predictive value of traditional histopathological features, attention has turned to the tumour microenvironment, particularly the stromal composition[13][14][15]. These studies have shown that the formation of dense, vascular stromal tissue along with angiogenesis is associated with worse outcomes, which may shield the tumour from the immune system or increase blood and nutrient flow. Mitotic scoring has been consistently shown to be an important risk predictor in DCIS’ invasive counterpart, invasive ductal carcinoma[16][17]. Though some work has been done to associate mitotic count with the Oncotype DX score [18], its relationship with disease progression in DCIS remains poorly explored. We seek to quantify cell types: lymphocytes, red blood cells (RBCs), fibroblasts, and mitotic cells; and associate their densities inside the tumour and stromal regions with eventual patient outcomes.

Modern machine learning techniques enable us to quickly analyse multiple features simultaneously. This allows us to develop more sophisticated risk scores and further investigate interactions among any set of features. It has the additional benefit of reducing interobserver variability in the annotation of digital pathology slides[19][20]. Using machine learning methods in this case allows us to conduct faster, more repeatable analysis over more slides. We hypothesise that AI systems capable of measuring the density of specific cell types in both tumour and stromal regions will be predictive of recurrence risk in DCIS. In this study, we use AI-based image analysis to quantify key stromal and epithelial cell populations in DCIS and assess their association with ipsilateral recurrence.

## 2 Methods

### 2.1 Breast Tissue Segmentation Models

Our method relies on accurate quantification of cell and tissue concentrations within a WSI. To accomplish this, we trained several models capable of segmenting cells and DCIS lesions for quantifying their densities across the entire slide.

#### 2.1.1 DCIS and Stroma Segmentation

To better understand the localisation of cells within the tumour microenvironment, we sought to develop a model capable of segmenting DCIS. We used a dataset previously described by Seth et al [21], which consists of 145 H&E WSIs derived from resection specimens of 145 women with DCIS. An expert pathologist reviewed each slide and marked the perimeter of all ducts containing DCIS. These segmentations were used to train a U-Net capable of recreating the pathologist’s labels. The resulting model had a dice score of 0.80. An example of the segmentation can be found in Figure 2.

To produce an annotation of the TAS region, we took a region around the perimeter of the lesion, including both the tumour boundary and the surrounding tissue. This was to ensure all stroma immediately adjacent to the tumour was accounted for in the analysis. An example of the resulting region is shown in Figure 1.

**Fig. 1.**
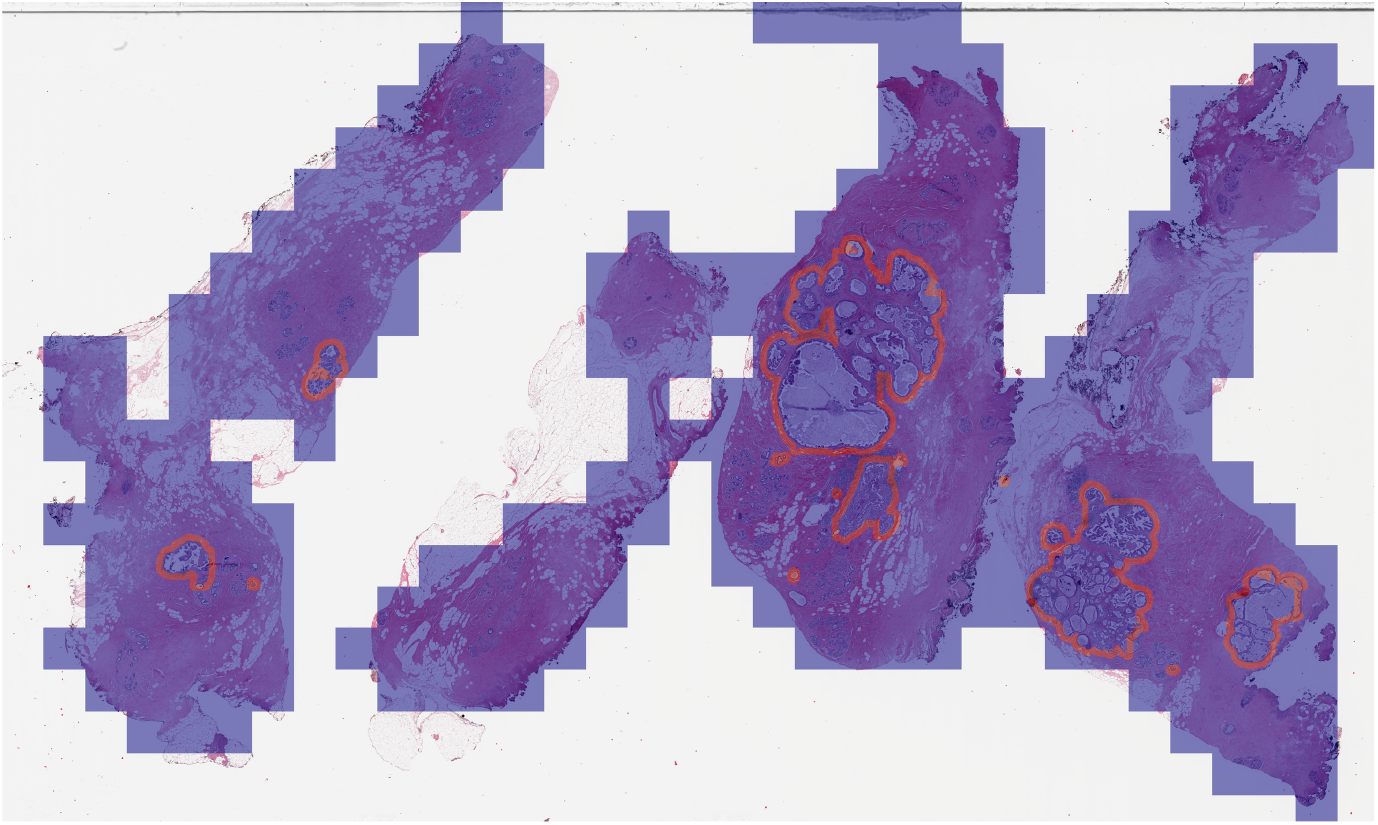
TAS (tumour-associated stroma) was defined as a 40-pixel buffer around DCIS ducts, including the tumour boundary and adjacent stroma. Red: TAS regions annotated based on the DCIS segmentation model. Blue: Slide area analysed by the DCIS model.

#### 2.1.2 Cellular Segmentation

We trained separate segmentation models for each of the cell types we are interested in. The cell types explored in this work are lymphocytes, mitotic cells, red blood cells, and cancer-associated fibroblasts and we obtained the annotated images required for training from several publicly available datasets.

For lymphocytes, we used a dataset assembled for the TIGER challenge[22]. This consisted of exact lymphocyte locations marked by a trained pathologist on excision slides from triple-negative and HER2+ invasive breast cancer patients. These were sourced from the TCGA-BRCA cohort (n=151), Radboud University Medical Center (n=26), and Jules Bordet Institute (n=18). For automatic quantification, we used our cell segmentation method developed in [23]. This uses a U-net architecture with a Resnet18 backbone to segment the area occupied by lymphocytes.

For mitotic segmentation, we used a dataset collected for the MIDOG22 challenge[24]. This took slides from various tissue sources, including human and canine tissue, and had a single pathologist label any mitotic figures in a given field of view. Three separate pathologists then evaluated each marked mitosis and marked it as either a true positive or a mimicker. Majority opinion was used to assign the final label. We used a variant of the model described by Jahanifar et al[25] to detect mitotic cells. Our final cell-type segmentations for red blood cells and fibroblasts came from the SegPath dataset[26]. This dataset consists of tissue sections stained initially with H&E and imaged. They were subsequently restained using immunohistochemistry (IHC) targeting proteins specific to the cell type of interest. In this case, *α*SMA was used to identify fibroblasts, and CD235a was used as the marker for red blood cells. The IHC images served as ground-truth labels in our experiments, and the U-Net was intended to reproduce their output solely from the H&E image. Examples of the resulting segmentations can be found in Figure 2. The three mitotic figures detected in the stromal/tumour boundary region, along with three examples of red blood cell clusters, are highlighted.

**Fig. 2.**
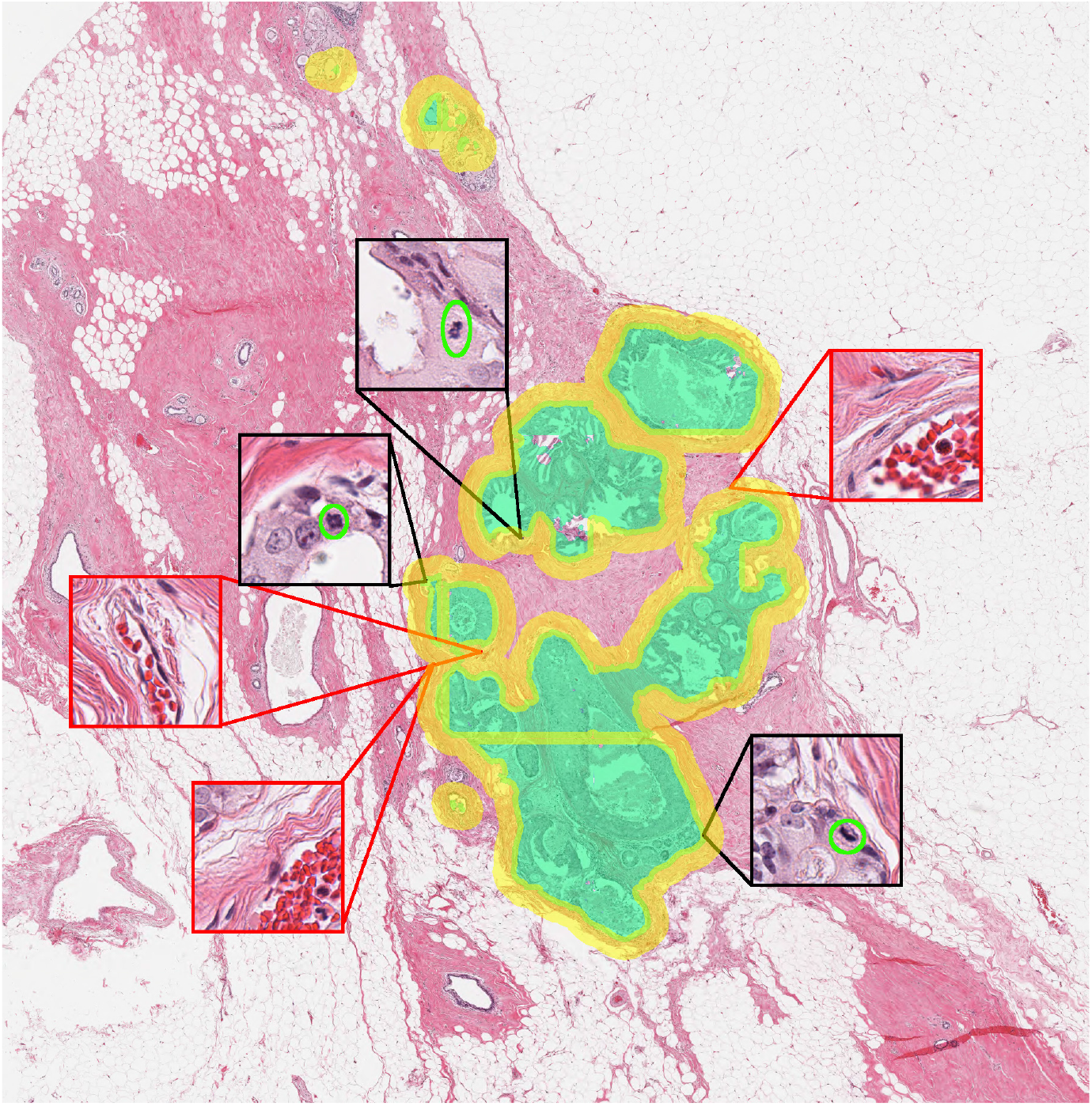
An example slide with DCIS labels (Green) and the automatically generated TAS labels (Yellow). Insets show examples of found mitotic figures (Border: Black) and clusters of RBCs (Border: Red).

### 2.2 DCIS Patient Risks Analysis

#### 2.2.1 Ontario DCIS Cohort

To determine the relative risk of different conditions in DCIS, we used the Ontario DCIS cohort[5]. In brief, this is a population-based cohort of women diagnosed with DCIS in Ontario between January 1994 and December 2003, assembled through linkage of provincial pathology and health databases, comprising 5,752 cases with pathology-verified DCIS. 3,762 of these women underwent breast-conserving surgery (BCS); and half receiving radiation treatment afterward. As described in Rakovitch et al[12], end-points were determined through linkage of Ontario Cancer Registry data, hospital records, and pathology reports. The primary endpoint was ipsilateral breast event (IBE), defined as any recurrence in the same breast—either invasive carcinoma or DCIS—after initial treatment. Our research focused on patients receiving only BCS, with the ultimate goal of identifying patients who will not have a recurrence without radiotherapy treatment. We were left with 1563 slides from 799 patients, with each patient having 1 or 2 slides in the dataset. Within this group, 165 patients had an IBE during the follow-up period, with 102 having an invasive recurrence and 63 having a recurrence of DCIS.

#### 2.2.2 Individual Predictor Hazard Ratios

We defined our features as the pixel-wise densities of our three cell types in both the tumour and TAS areas. We further tested whether the number of separate DCIS foci on the slide or the relative percentage of DCIS tissue to total tissue correlated with patient prognosis. This left us with a total of eight features to test. For patients with multiple slides, we averaged the densities across both to give a single value. Features which were particularly concentrated around 0, being RBC, mitotic, and lesion density, were transformed with a logit function to match a normal distribution better. All variables were then normalised across the entire dataset to have a mean of 0 and a standard deviation of 1. The Python Lifelines package[27] was used to calculate the hazard ratios of the variables, also calculating the p-value using the Wald test. These were adjusted using the Benjamini-Hochberg (BH) method [28] to re-evaluate the false discovery rate across the multiple tests.

To analyse non-linear associations with outcome, we investigated the effectiveness of binning patients rather than treating them continuously. To divide patients into groups with low or high concentrations, we used a 2-class Gaussian mixture model trained on the entire dataset. This was to create separation in an unbiased way.

#### 2.2.3 Patient Stromal Type Clustering

The densities of several different cell types in the peritumoral region were shown to associate with outcomes. A second GMM was used to cluster patients into discrete phenotypic groups based on their cell-stromal densities, specifically using the mitosis-TAS, lymphocyte-TAS, RBC-TAS, and number of lesions variables. The number of clusters was chosen to optimise the Bayesian information criterion. The mean of each feature across clusters was reported, and hazard ratios between clusters were calculated. Cluster 1 was arbitrarily selected as the reference for the hazard ratios.

## 3 Results

### 3.1 Continuous Variable Investigation

The investigation of the continuous variables showed that some features carried prognostic value Figure 3. The number of foci rather than the overall DCIS area was shown to have a stronger association with patient outcome. Additionally, mitotic density in the TAS region was found to have a worse prognosis than density inside the DCIS region itself. RBC density in the stroma and the tumour region were shown to have a relatively weak, but notable association with poorer patient outcomes.

**Fig. 3.**
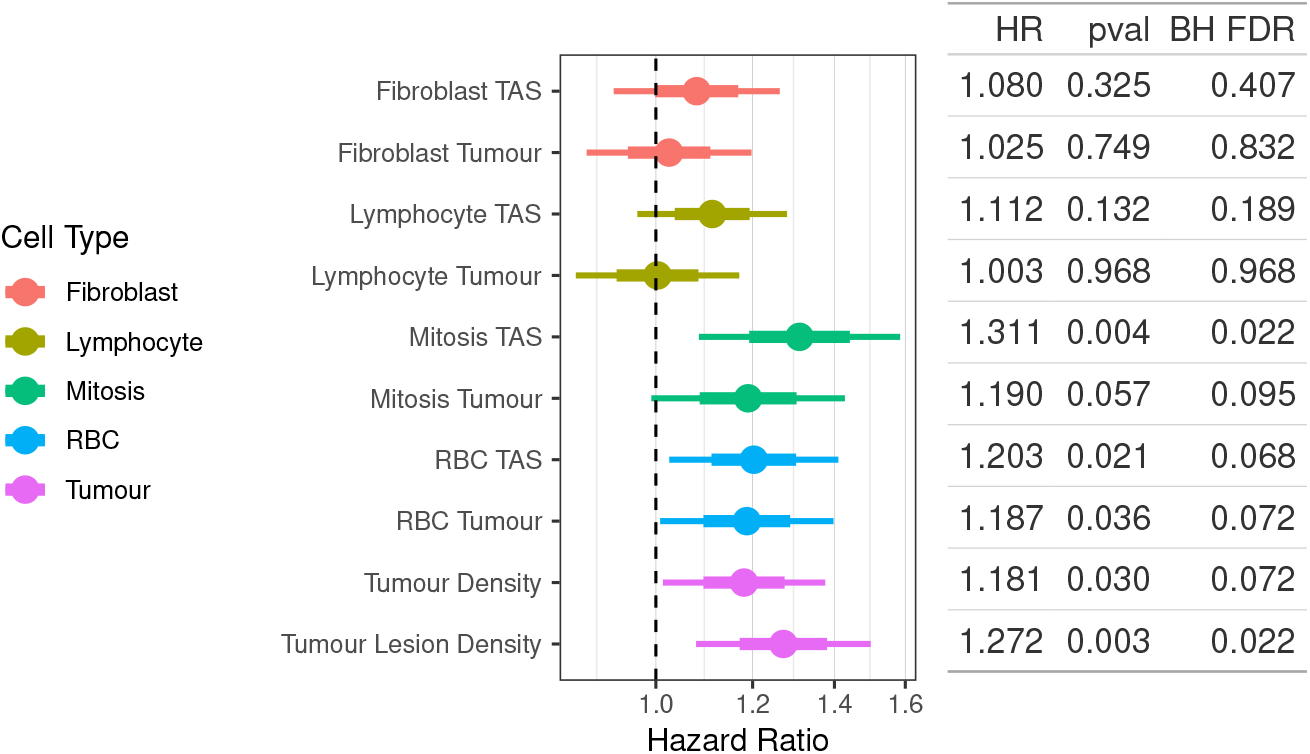
Plot represents the univariate Cox model coefficients for each feature generated for the segmentation models. Since all are normalised to have a standard deviation of 1, this coefficient represents the hazard ratio (HR) for a 1-standard-deviation increase in the feature. The table shows the associated p-value and the Benjamini-Hochberg (BH) adjusted false discovery rate (FDR).

No prognostic value was detected in this assay for either the lymphocyte or fibroblast density.

### 3.2 Binarized Feature Evaluation

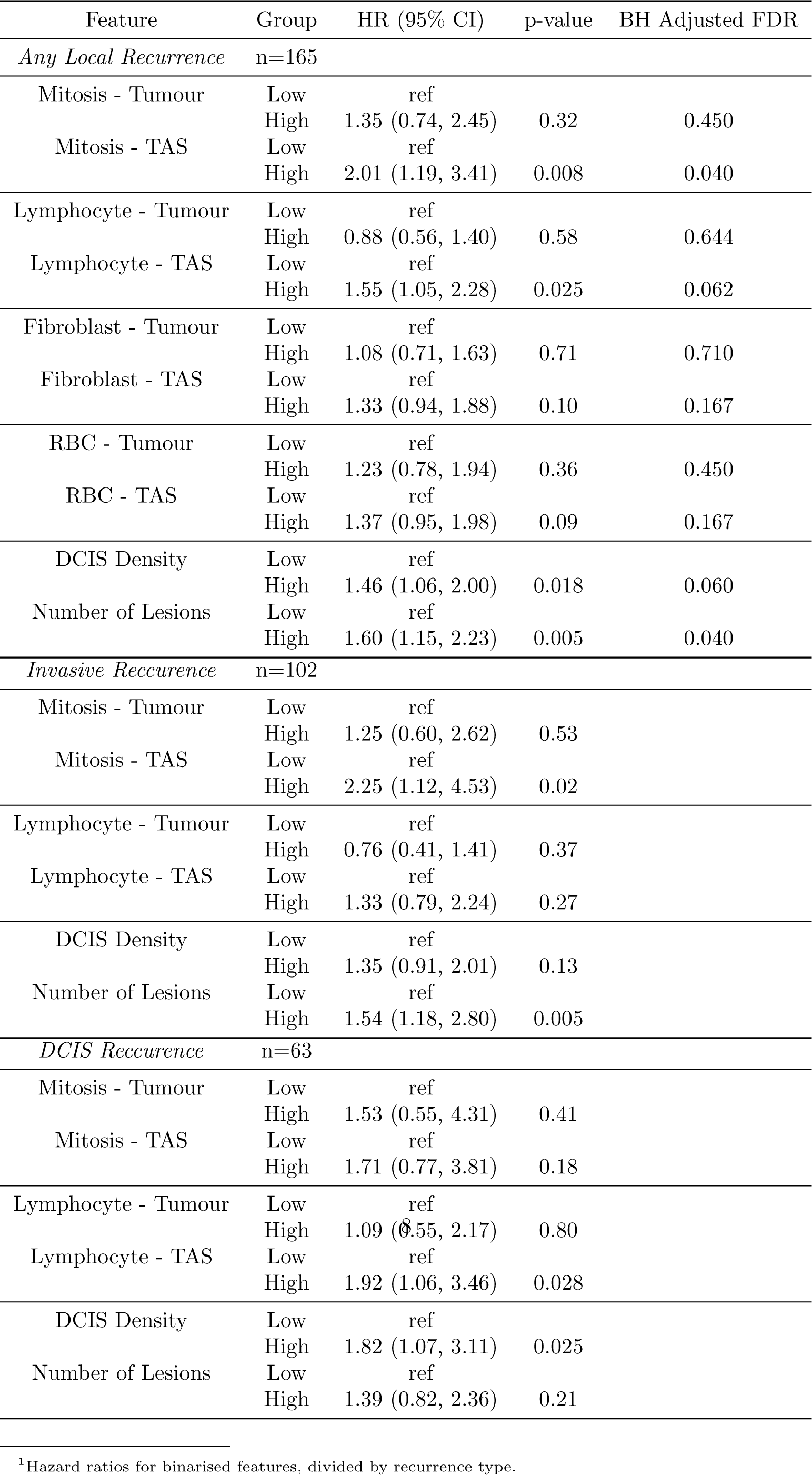

In contrast to the investigation of continuous variables, a small signal was detected in the lymphocyte-TAS density after binarisation was carried out, with higher lymphocyte concentration associated with worse outcomes. This is in line with previous findings[29][30].

Higher mitotic rates in the TAS and the number of lesions were again shown to have the strongest associations with outcomes. In contrast to lymphocytes, RBC densities showed no strong association after binarisation, suggesting that treating this feature as a continuous, linear variable is more appropriate.

### 3.3 Patient Clustering on Stromal Features

Cellular density in the stromal tissue surrounding DCIS lesions was indicated to have predictive power for patient outcome in three cell types investigated here: mitotic cells, lymphocytes, and red blood cells. We combined these, along with our most important feature, the density of distinct lesions, into clusters of patients using a GMM. We optimised the number of clusters by minimising the Bayesian information criterion (BIC). This left us with four stromal patient phenotypes (Figure 4).

**Fig. 4.**
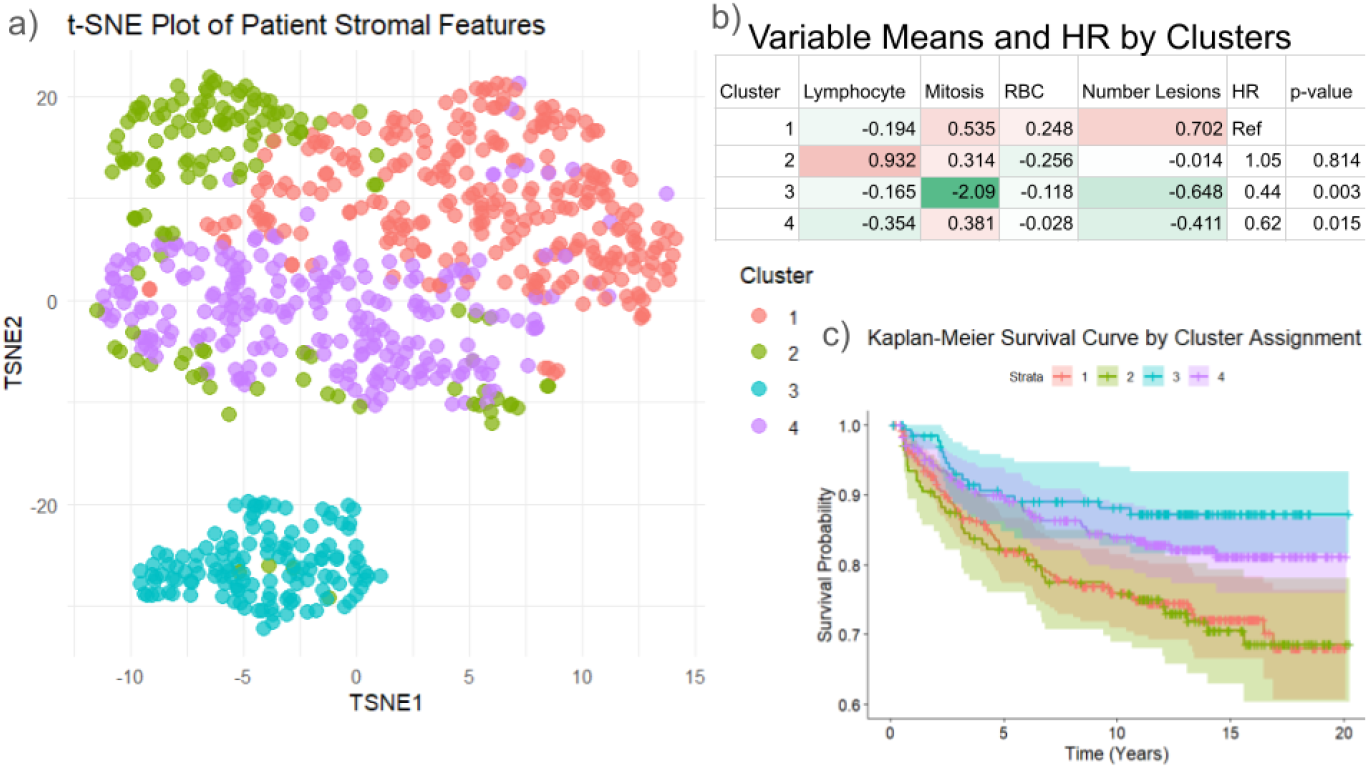
a) t-SNE map of patient stromal features, colored by their cluster assignments from the GMM. We notice a very distinct clustering for cluster 3 compared to the other groups; this is due to the large separation between patients with no mitoses and those with any. b) Table listing the mean value of each feature and the hazard ratio between each cluster and its p-value. Variable values were normalised for the entire population to have a mean of 0 and a standard deviation of 1 before clustering. This means that a cluster mean of 1 represents a cluster whose mean is 1 is a single standard deviation greater than the whole population. c) Kaplan-Meier survival curve for each cluster.

Phenotype 1 shows a high concentration of mitotic cells and individual lesions, while phenotype 2 shows a high density of lymphocytes and a more typical density of other features. Both of these groups have very similar risk profiles. This may indicate that there are two subtypes of high-risk DCIS tumours. Similarly, phenotypes 3 and 4 also have very similar risk profiles. Phenotype 3 is dominated mainly by patients with extremely low mitotic rates (essentially no mitoses found) and fewer lesions. Group 4 has the lowest immune concentration found, along with a low concentration of lesions. There is a greater diference in the risk profiles of groups 3 and 4 compared with the two high risk groups but this is not statistically significant (HR=1.42, p-value=0.22).

## 4 Discussion

In this work, we demonstrated the potential of various stromal features for predicting local recurrence in DCIS. This is consistent with previous work, which showed that lymphocyte density in the surrounding region rather than the tumour itself is an indicator of poor prognosis in DCIS[29][30]. We have found that this relationship is potentially non-linear; future work with larger datasets should investigate other non-linear models to see whether they improve predictive power.

The number of individual DCIS ducts present in the specimen and the proportion of DCIS tissue relative to total tissue area were both associated with recurrence. Notably, the duct count provided slightly stronger predictive value. This modest improvement may suggest it is indicating something similar to multifocality, which has also been shown to be a biomarker of recurrence risk in DCIS[31]. In addition, both size-related measures may function as surrogates for larger-scale pathological characteristics, such as the lesion approaching a palpable size, which is typically associated with more aggressive oncological features[32].

There was only a weak association between mitotic activity within the tumour bed and patient outcome, but a much stronger association emerged when mitoses were assessed in the TAS region, which in our case was defined to include the tumour edge.

This aligns with practice in invasive tumours, where mitotic hotspots are typically sought at the tumour periphery[33]. However, several studies have emphasized the need for more rigorous and standardized hotspot selection to improve measurement reproducibility[34][35]. Future work could examine whether focusing on predefined regions, such as the periphery in this study, yields more informative scores than relying solely on visually selected hotspots.

Red blood cell density in the peritumoral regions was found to correlate negatively with patient outcomes. Even low-proliferation breast cancer types such as ductal carcinoma in situ (DCIS) are known to initiate vascular remodelling and angiogenesis [36][14]. Increased vascular density, particularly microvascular density, has been associated with elevated risks of malignancy and recurrence across various cancer types [37][38][39]. In these works, vascular density is quantified using immunohistochemical (IHC) staining for endothelial markers, followed by manual counting of vessel-like structures by human observers. We hypothesise that, in our case, RBC density serves as a crude surrogate for vascular density. The SegPath dataset includes paired H&E and IHC-stained images for endothelial cells. It may be beneficial to develop a model capable of segmenting endothelial cells from H&E images, thereby eliminating the need for a specialised stain for quantification. This may work in combination with our existing RBC and fibroblast models to provide a more comprehensive view of the vascular landscape and enable quantification of extravascular RBCs, potentially characterising a microhemorrhagic environment that contributes to tumour progression and poor prognosis[40].

Although an increasing body of work has demonstrated that fibroblasts play a significant role in DCIS progression[41][42][43], we found no association with patient outcome. This could be due to the lack of specificity of our protein marker. Our approach relies on segmenting cells which were tagged with *α*SMA IHC. Though this population is consistently linked to cancer-associated fibroblasts[44], previous DCIS studies have used other markers, such as Cav-1 as a marker for healthy fibroblasts[45][41] or PDGFR for CAFs[43]. It is possible that a more targeted fibroblast model, trained using labels derived from a more specific IHC protein would yield more informative results.

Our investigation into creating patient stromal subtypes showed promise in identifying patient risk groups. Two distinct high-risk groups were identified, one with lesions with more features directly associated with proliferation and a second with lesions that featured a greater immune presence. While this stratification should be viewed as exploratory, since the variables were selected using the same dataset, it nonetheless suggests that deeper investigation into the stromal composition of tumours may enhance treatment decision-making. These findings warrant further investigations in larger cohorts, with held-out test sets. This would allow for the future development of validated risk scores, helping correctly identify patients who need radiotherapy and those who can forgo it.

Our findings suggest that stromal composition may provide prognostic information for DCIS. Integration of these AI-derived biomarkers with existing tools such as clinical nomograms could enhance individualised treatment planning, potentially reducing overtreatment while maintaining oncologic safety. However, this study has limitations, notably its retrospective design, reliance on a single population-based cohort, and lack of external validation. Furthermore, model development and feature selection were performed on the same dataset, potentially overestimating predictive performance. Future work should focus on prospective validation in multi-institutional cohorts, exploration of non-linear modelling approaches, and incorporation of stromal phenotypes into clinically actionable risk scores. Ultimately, these efforts could support precision medicine strategies that identify patients who can safely omit adjuvant radiotherapy after breast-conserving surgery.

## Data Availability

All data produced in the present study are available upon reasonable request to the authors

## Supplementary information

To train our cell segmentation model on mitosis, fibroblast detection, and red blood cells, we used a U-Net with an EfficientNet [46] backbone, we used a deeper model than the MIDOG winner (B7 vs. B0) and eschewed the secondary classification head. We found this maintained accuracy while prioritising speed, which is necessary to apply it across an entire WSI. The training data for this challenge features 50 high-powered fields from 3 different scanners. Mitotic figures were initially labelled as bounding boxes; to convert to segmentation, a 19-pixel-radius circle was used as the ground truth in the centre of the provided bounding box. The images and corresponding masks were tiled to produce 512×512 pixel patches. During training, patches containing mitosis were oversampled so that each batch contained roughly equal numbers of positive and negative patches. To improve generalizability, we found greater success using a RandAugment scheme[47] rather than stain normalisation. Training the DCIS segmentation model was similar, this time taking patches at 2.0mpp. 115 slides were used for training and 30 for validation. To produce an annotation of the boundary TAS region, we used a 40-pixel buffer at 2.0 microns per pixel magnification as a definition of the stromal region surrounding each duct. This comprised the 10-pixel-diameter region inside the tumour annotation and a 30-pixel-diameter region outside.

## Acknowledgements

This work was supported by funding from the Canadian Institutes of Health Research (CIHR), the Canadian Breast Cancer Foundation (CBCF), the Canada Foundation for Innovation and the Ontario Research Fund. We thank Shazia Akbar and Sabina Trebinjac for their contributions to data curation, which were essential to the completion of this study.

This study made use of data from the ICES Data Repository, which is managed by the Institute for Clinical Evaluative Sciences with support from its funders and partners: Canada’s Strategy for Patient-Oriented Research (SPOR), the Ontario SPOR Support Unit, the Canadian Institutes of Health Research and the Government of Ontario. The opinions, results and conclusions reported are those of the authors. No endorsement by ICES or any of its funders or partners is intended or should be inferred. Parts of this material are based on data and information compiled and provided by CIHI. However, the analyses, conclusions, opinions and statements expressed herein are those of the author, and not necessarily those of CIHI.

## References

[1] Ernster, V.L., Ballard-Barbash, R., Barlow, W.E., Zheng, Y., Weaver, D.L., Cutter, G., Yankaskas, B.C., Rosenberg, R., Carney, P.A., Kerlikowske, K., et al.: Detection of ductal carcinoma in situ in women undergoing screening mammography. Journal of the National Cancer Institute 94(20), 1546–1554 (2002)

[2] Fisher, B., Land, S., Mamounas, E., Dignam, J., Fisher, E.R., Wolmark, N.: Prevention of invasive breast cancer in women with ductal carcinoma in situ: an update of the national surgical adjuvant breast and bowel project experience. In: Seminars in Oncology, vol. 28, pp. 400–418 (2001). Elsevier

[3] Bijker, N., Meijnen, P., Peterse, J.L., Bogaerts, J., Van Hoorebeeck, I., Julien, J.-p., Gennaro, M., Rouanet, P., Avril, A., Fentiman, I.S., et al.: Breast-conserving treatment with or without radiotherapy in ductal carcinoma-in-situ: ten-year results of european organisation for research and treatment of cancer randomized phase iii trial 10853—a study by the eortc breast cancer cooperative group and eortc radiotherapy group. Journal of clinical oncology 24(21), 3381–3387 (2006)

[4] Holmberg, L., Garmo, H., Granstrand, B., Ringberg, A., Arnesson, L.-G., Sandelin, K., Karlsson, P., Anderson, H., Emdin, S.: Absolute risk reductions for local recurrence after postoperative radiotherapy after sector resection for ductal carcinoma in situ of the breast. Journal of Clinical Oncology 26(8), 1247–1252 (2008)

[5] Rakovitch, E., Nofech-Mozes, S., Narod, S.A., Hanna, W., Thiruchelvam, D., Saskin, R., Taylor, C., Tuck, A., Sengupta, S., Elavathil, L., et al.: Can we select individuals with low risk ductal carcinoma in situ (dcis)? a population-based outcomes analysis. Breast cancer research and treatment 138(2), 581–590 (2013)

[6] Cuzick, J., Sestak, I., Pinder, S.E., Ellis, I.O., Forsyth, S., Bundred, N.J., Forbes, J.F., Bishop, H., Fentiman, I.S., George, W.D.: Effect of tamoxifen and radio-therapy in women with locally excised ductal carcinoma in situ: long-term results from the uk/anz dcis trial. The lancet oncology 12(1), 21–29 (2011)

[7] Francis, A., Thomas, J., Fallowfield, L., Wallis, M., Bartlett, J.M., Brookes, C., Roberts, T., Pirrie, S., Gaunt, C., Young, J., et al.: Addressing overtreatment of screen detected dcis; the loris trial. European journal of cancer 51(16), 2296–2303 (2015)

[8] Groen, E.J., Elshof, L.E., Visser, L.L., Rutgers, E.J.T., Winter-Warnars, H.A., Lips, E.H., Wesseling, J.: Finding the balance between over-and under-treatment of ductal carcinoma in situ (dcis). The Breast 31, 274–283 (2017)

[9] Rudloff, U., Jacks, L.M., Goldberg, J.I., Wynveen, C.A., Brogi, E., Patil, S., Van Zee, K.J.: Nomogram for predicting the risk of local recurrence after breast-conserving surgery for ductal carcinoma in situ. Journal of clinical oncology 28(23), 3762–3769 (2010)

[10] Wang, S.-Y., Shamliyan, T., Virnig, B.A., Kane, R.: Tumor characteristics as predictors of local recurrence after treatment of ductal carcinoma in situ: a meta-analysis. Breast cancer research and treatment 127(1), 1–14 (2011)

[11] Zhang, X., Dai, H., Liu, B., Song, F., Chen, K.: Predictors for local invasive recurrence of ductal carcinoma in situ of the breast: a meta-analysis. European Journal of Cancer Prevention 25(1), 19–28 (2016)

[12] Rakovitch, E., Nofech-Mozes, S., Hanna, W., Baehner, F.L., Saskin, R., Butler, S.M., Tuck, A., Sengupta, S., Elavathil, L., Jani, P.A., et al.: A population-based validation study of the dcis score predicting recurrence risk in individuals treated by breast-conserving surgery alone. Breast cancer research and treatment 152(2), 389–398 (2015)

[13] Brown, L.F., Guidi, A.J., Schnitt, S.J., Van De Water, L., Iruela-Arispe, M.L., Yeo, T.-K., Tognazzi, K., Dvorak, H.F.: Vascular stroma formation in carcinoma in situ, invasive carcinoma, and metastatic carcinoma of the breast. Clinical Cancer Research 5(5), 1041–1056 (1999)

[14] Pavlakis, K., Messini, I., Vrekoussis, T., Yiannou, P., Keramopoullos, D., Louvrou, N., Liakakos, T., Stathopoulos, E.N.: The assessment of angiogenesis and fibroblastic stromagenesis in hyperplastic and pre-invasive breast lesions. BMC cancer 8(1), 88 (2008)

[15] Risom, T., Glass, D.R., Averbukh, I., Liu, C.C., Baranski, A., Kagel, A., McCaffrey, E.F., Greenwald, N.F., Rivero-Gutiérrez, B., Strand, S.H., et al.: Transition to invasive breast cancer is associated with progressive changes in the structure and composition of tumor stroma. Cell 185(2), 299–310 (2022)

[16] Bloom, H., Richardson, W.: Histological grading and prognosis in breast cancer: a study of 1409 cases of which 359 have been followed for 15 years. British journal of cancer 11(3), 359 (1957)

[17] Chang, J.M., McCullough, A.E., Dueck, A.C., Kosiorek, H.E., Ocal, I.T., Lidner, T.K., Gray, R.J., Wasif, N., Northfelt, D.W., Anderson, K.S., et al.: Back to basics: traditional nottingham grade mitotic counts alone are significant in predicting survival in invasive breast carcinoma. Annals of surgical oncology 22(Suppl 3), 509–515 (2015)

[18] Knopfelmacher, A., Fox, J., Lo, Y., Shapiro, N., Fineberg, S.: Correlation of histopathologic features of ductal carcinoma in situ of the breast with the oncotype dx dcis score. Modern Pathology 28(9), 1167–1173 (2015)

[19] Veta, M., Van Diest, P.J., Jiwa, M., Al-Janabi, S., Pluim, J.P.: Mitosis counting in breast cancer: Object-level interobserver agreement and comparison to an automatic method. PloS one 11(8), 0161286 (2016)

[20] Rabe, K., Snir, O.L., Bossuyt, V., Harigopal, M., Celli, R., Reisenbichler, E.S.: Interobserver variability in breast carcinoma grading results in prognostic stage differences. Human pathology 94, 51–57 (2019)

[21] Seth, N., Akbar, S., Nofech-Mozes, S., Salama, S., Martel, A.L.: Automated segmentation of dcis in whole slide images. In: European Congress on Digital Pathology, pp. 67–74 (2019). Springer

[22] Rijthoven, M., Aswolinskiy, W., Tessier, L., Balkenhol, M., Bogaerts, J.M., Drubay, D., Blesa, L.C., Peeters, D., Stovgaard, E.S., Lænkholm, A.-V., et al.: Tumor-infiltrating lymphocytes in breast cancer through artificial intelligence: biomarker analysis from the results of the tiger challenge. medRxiv, 2025–02 (2025)

[23] Ramanathan, V., Pati, P., McNeil, M., Martel, A.L.: Ensemble of prior-guided expert graph models for survival prediction in digital pathology. In: International Conference on Medical Image Computing and Computer-Assisted Intervention, pp. 262–272 (2024). Springer

[24] Aubreville, M., Stathonikos, N., Donovan, T.A., Klopfleisch, R., Ammeling, J., Ganz, J., Wilm, F., Veta, M., Jabari, S., Eckstein, M., et al.: Domain generalization across tumor types, laboratories, and species—insights from the 2022 edition of the mitosis domain generalization challenge. Medical Image Analysis 94, 103155 (2024)

[25] Jahanifar, M., Shephard, A., Zamanitajeddin, N., Graham, S., Raza, S.E.A., Minhas, F., Rajpoot, N.: Mitosis detection, fast and slow: Robust and efficient detection of mitotic figures. Medical Image Analysis 94, 103132 (2024)

[26] Komura, D., Onoyama, T., Shinbo, K., Odaka, H., Hayakawa, M., Ochi, M., Herdiantoputri, R.R., Endo, H., Katoh, H., Ikeda, T., et al.: Restaining-based annotation for cancer histology segmentation to overcome annotation-related limitations among pathologists. Patterns 4(2) (2023)

[27] Davidson-Pilon, C.: lifelines: survival analysis in Python. https://github.com/CamDavidsonPilon/lifelines. Version 0.27.7 (2023)

[28] Benjamini, Y., Hochberg, Y.: Controlling the false discovery rate: A practical and powerful approach to multiple testing. Journal of the Royal Statistical Society: Series B (Methodological) 57(1), 289–300 (1995)

[29] Schiza, A., Thurfjell, V., Tullberg, A.S., Olofsson, H., Lindberg, A., Holmberg, E., Bremer, T., Micke, P., Karlsson, P., Wärnberg, F., et al.: Tumour-infiltrating lymphocytes add prognostic information for patients with low-risk dcis: findings from the swedcis randomised radiotherapy trial. European Journal of Cancer 168, 128–137 (2022)

[30] Li, H., Aggarwal, A., Toro, P., Fu, P., Badve, S.S., Cuzick, J., Madabhushi, A., Thorat, M.A.: A prognostic and predictive computational pathology immune signature for ductal carcinoma in situ: retrospective results from a cohort within the uk/anz dcis trial. The Lancet Digital Health 6(8), 562–569 (2024)

[31] Rakovitch, E., Pignol, J.-P., Hanna, W., Narod, S., Spayne, J., Nofech-Mozes, S., Chartier, C., Paszat, L.: Significance of multifocality in ductal carcinoma in situ: outcomes of women treated with breast-conserving therapy. Journal of clinical oncology 25(35), 5591–5596 (2007)

[32] Rajan, S.S., Verma, R., Shaaban, A.M., Sharma, N., Dall, B., Lansdown, M.: Palpable ductal carcinoma in situ: analysis of radiological and histological features of a large series with 5-year follow-up. Clinical Breast Cancer 13(6), 486–491 (2013)

[33] Ibrahim, A., Lashen, A.G., Katayama, A., Mihai, R., Ball, G., Toss, M.S., Rakha, E.A.: Defining the area of mitoses counting in invasive breast cancer using whole slide image. Modern Pathology 35(6), 739–748 (2022)

[34] Bonert, M., Tate, A.J.: Mitotic counts in breast cancer should be standardized with a uniform sample area. Biomedical engineering online 16(1), 28 (2017)

[35] Rakha, E.A., Bennett, R.L., Coleman, D., Pinder, S.E., Ellis, I.O.: Review of the national external quality assessment (eqa) scheme for breast pathology in the uk. Journal of clinical pathology 70(1), 51–57 (2017)

[36] Lee, A.H., Happerfield, L.C., Bobrow, L.G., Millis, R.R.: Angiogenesis and inflammation in ductal carcinoma in situ of the breast. The Journal of Pathology: A Journal of the Pathological Society of Great Britain and Ireland 181(2), 200–206 (1997)

[37] Weidner, N., Semple, J.P., Welch, W.R., Folkman, J.: Tumor angiogenesis and metastasis—correlation in invasive breast carcinoma. New England Journal of Medicine 324(1), 1–8 (1991)

[38] Guidi, A.J., Fischer, L., Harris, J.R., Schnitt, S.J.: Microvessel density and distribution in ductal carcinoma in situ of the breast. JNCI: Journal of the National Cancer Institute 86(8), 614–619 (1994)

[39] Uzzan, B., Nicolas, P., Cucherat, M., Perret, G.-Y.: Microvessel density as a prognostic factor in women with breast cancer: a systematic review of the literature and meta-analysis. Cancer research 64(9), 2941–2955 (2004)

[40] Yin, T., He, S., Liu, X., Jiang, W., Ye, T., Lin, Z., Sang, Y., Su, C., Wan, Y., Shen, G., et al.: Extravascular red blood cells and hemoglobin promote tumor growth and therapeutic resistance as endogenous danger signals. The Journal of Immunology 194(1), 429–437 (2015)

[41] Martinez-Outschoorn, U.E., Pavlides, S., Whitaker-Menezes, D., Daumer, K.M., Milliman, J.N., Chiavarina, B., Migneco, G., Witkiewicz, A.K., Martinez-Cantarin, M.P., Flomenberg, N., et al.: Tumor cells induce the cancer associated fibroblast phenotype via caveolin-1 degradation: implications for breast cancer and dcis therapy with autophagy inhibitors. Cell Cycle 9(12), 2423–2433 (2010)

[42] Erez, N., Glanz, S., Raz, Y., Avivi, C., Barshack, I.: Cancer associated fibroblasts express pro-inflammatory factors in human breast and ovarian tumors. Biochemical and biophysical research communications 437(3), 397–402 (2013)

[43] Strell, C., Paulsson, J., Jin, S.-B., Tobin, N.P., Mezheyeuski, A., Roswall, P., Mutgan, C., Mitsios, N., Johansson, H., Wickberg, S.M., et al.: Impact of epithelial–stromal interactions on peritumoral fibroblasts in ductal carcinoma in situ. JNCI: Journal of the National Cancer Institute 111(9), 983–995 (2019)

[44] Nurmik, M., Ullmann, P., Rodriguez, F., Haan, S., Letellier, E.: In search of definitions: Cancer-associated fibroblasts and their markers. International journal of cancer 146(4), 895–905 (2020)

[45] Witkiewicz, A.K., Dasgupta, A., Nguyen, K., Liu, C., Kovatich, A.J., Schwartz, G.F., Pestell, R.G., Sotgia, F., Rui, H., Lisanti, M.P.: Stromal caveolin-1 levels predict early dcis progression to invasive breast cancer. Cancer biology & therapy 8(11), 1071–1079 (2009)

[46] Tan, M., Le, Q.: Efficientnet: Rethinking model scaling for convolutional neural networks. In: International Conference on Machine Learning, pp. 6105–6114 (2019). PMLR

[47] Cubuk, E.D., Zoph, B., Shlens, J., Le, Q.V.: Randaugment: Practical automated data augmentation with a reduced search space. In: Proceedings of the IEEE/CVF Conference on Computer Vision and Pattern Recognition Workshops, pp. 702–703 (2020)

